# Mitochondrial Bioenergetics in Resilience of Older Adults with Gynecologic Cancer: Design and Rationale of a Pilot Study

**DOI:** 10.1101/2025.06.02.25328174

**Authors:** Anna Kuan-Celarier, Michelle L. Wallander, Jennifer Hartzell, Brittany Lees, Xiaoyan Iris Leng, Philip A. Kramer, Nicholas J. Day, Wei-Jun Qian, Bumsoo Ahn

## Abstract

Resilience, the ability to recover and maintain function following stresses, is a critical factor influencing treatment tolerance and recovery in older adults with cancer. Despite the high incidence of gynecologic cancers in postmenopausal individuals, resilience in this population remains underexplored, even though patients commonly face compounded stress from both chemotherapy and surgery. The goal of our research is 1) to test the feasibility of cognitive and physical function assessments in older women with gynecologic cancers and 2) to discover reliable predictors that enhance clinical decision-making and guide personalized treatment strategies. Current clinical assessments focus on isolated physiological systems. As such, there is a need for a reliable predictor that captures systemic resilience more comprehensively. A reliable predictor of resilience following cancer treatment could improve clinical decision-making and identify potential targets for therapeutic intervention. Both mitochondrial bioenergetics and oxidative stress are presumably mechanistically linked to resilience of patients with gynecologic cancers because of widely known effects of chemotherapy and tumor burden on mitochondrial bioenergetics. Mitochondria generate more than 95% of cellular ATP through oxidative phosphorylation, a process essential for recovery following physiological stress. Oxidative stress disrupts excitation–contraction coupling and reduces metabolic efficiency in skeletal muscle, contributing to weakness and fatigue. In the brain, oxidative modifications have been associated with impaired neurotransmission and cognitive dysfunction. This protocol paper describes a longitudinal study design aimed at evaluating the feasibility of resilience assessment and testing mitochondria and oxidative stress as predictors of resilience in older adults diagnosed with advanced endometrial or ovarian cancer.

## Introduction

Resilience—the capacity to recover and maintain function following physiological or psychological stress (Whitson et al., 2016)—is a critical determinant of how older adults tolerate cancer treatments (Walston et al., 2023; Whitson et al., 2016). While cancer incidence exponentially rises with age (Rozhok and DeGregori, 2019) and gynecologic cancers are predominantly diagnosed post-menopause (Surveillance et al., 2024a; Surveillance et al., 2024b; Surveillance et al., 2024c), little is known about the resilience of this population, particularly in the context of both physical and cognitive function. Older patients with gynecologic cancer must endure not only the burden of the tumor but also the physiological demands of chemotherapy and cytoreductive surgery, often compounded by age-related decline (Duan-Porter et al., 2016; Walston et al., 2023). However, studying resilience in this population poses practical challenges: treatment-related fatigue, transportation limitations, and comorbidities may hinder research participation. Moreover, resilience is a relatively novel construct in oncology, and few studies have jointly examined physical and cognitive resilience in this setting. The primary objectives of our pilot study are to test the feasibility of assessing both physical and cognitive function longitudinally in older women undergoing standard of care treatment for gynecologic cancers, and to test promising predictors of resilience which can help guide clinical decision-making.

Several stress tests have been proposed as predictors of resilience across different patient populations. Examples of these molecular stress tests include adrenocorticotropic hormone response, the glucose tolerance testing (Walston et al., 2023), and measurement of cytokine levels and endotoxins from whole blood (Mehta et al., 2012). While these tests focus on isolated physiological systems, such as metabolic or immune responses, there is an urgent need for a predictor that captures systemic resilience more comprehensively. A reliable predictor of resilience after cancer treatment could enhance clinical decision-making and help identify potential targets for therapeutic intervention.

Mitochondria are recognized as key contributors to physical and cognitive resilience (Colon-Emeric et al., 2025) due to their role in energy production for high-demand organs such as muscles and the brain. Mitochondria produce greater than 95% of cellular ATP through oxidative phosphorylation (Tzameli, 2012). Energy in the form of ATP serves as the fuel for cellular activity; ATP deficiency due to mitochondrial dysfunction decreases tolerance of chemotherapy and surgical stress (Jatoi et al., 2000). Further evidence of mitochondrial dysfunction, such as impaired cellular membrane potential and increased oxidative stress, has also been seen in older adults and patients undergoing cancer treatment (Cauli, 2021; Zandbergen et al., 2019). To our knowledge, the relationship between mitochondrial bioenergetics and physical and cognitive resilience in older patients with cancer has not been investigated.

Reactive oxygen species (ROS)—primarily released during mitochondrial electron transport systemactivity—is a major contributor to functional decline associated with aging and chronic disease, impairing both cognitive and physical performance. At the molecular level, ROS can disrupt cellular homeostasis by damaging proteins, lipids, and nucleic acids (Sies and Jones, 2020). Among the most sensitive targets of oxidative stress are cysteine residues in proteins, which undergo reversible and irreversible oxidative modifications that alter protein structure, localization, and function (Lennicke and Cochemé, 2021). This redox-sensitive regulation is particularly important in signaling pathways related to mitochondrial function, synaptic plasticity, and muscle contractility. In skeletal muscle, cysteine oxidation disrupts excitation– contraction coupling and metabolic efficiency, contributing to weakness and fatigue (Bruton et al., 2008; Dutka et al., 2012; Kramer et al., 2015), whereas in the brain, oxidative modifications have been linked to impaired neurotransmission and cognitive dysfunction (Cobley et al., 2018). Given the heightened oxidative stress observed in patients undergoing chemotherapy, understanding the extent and impact of cysteine oxidation may reveal critical mechanisms underlying diminished physical and cognitive resilience in older cancer patients.

The primary goal of our pilot study is to determine the feasibility of assessing resilience longitudinally in older adults with newly diagnosed advanced endometrial or ovarian cancer. Additionally, the study will test whether mitochondrial respiration and oxidative stress predict changes in cognitive and physical resilience of patients with gynecologic cancer receiving cancer treatments. By performing these analyses, our research seeks to uncover biologic factors contributing to resilience, potentially leading to innovative strategies to enhance resilience. Enhanced understanding of cognitive and physical resilience can help identify ways to improve patient outcomes and preserve function and quality of life for older patients with cancer.

## Methods

### Study design, setting, and eligibility criteria

This prospective clinical study is being conducted at Atrium Health Wake Forest Baptist Comprehensive Cancer Center (AHWFBCCC). Participants are enrolled at two campuses located in Winston-Salem and Charlotte, North Carolina. Study coordinators pre-screen appropriate patients to assess eligibility before their first appointment with an oncologist. All participants provide informed consent. The overall study design is illustrated in Fig. 1. This protocol was approved by the Wake Forest School of Medicine Institutional Review Board. The study is funded by the AHWFBCCC and the American Cancer Society.

**Fig. 1.**
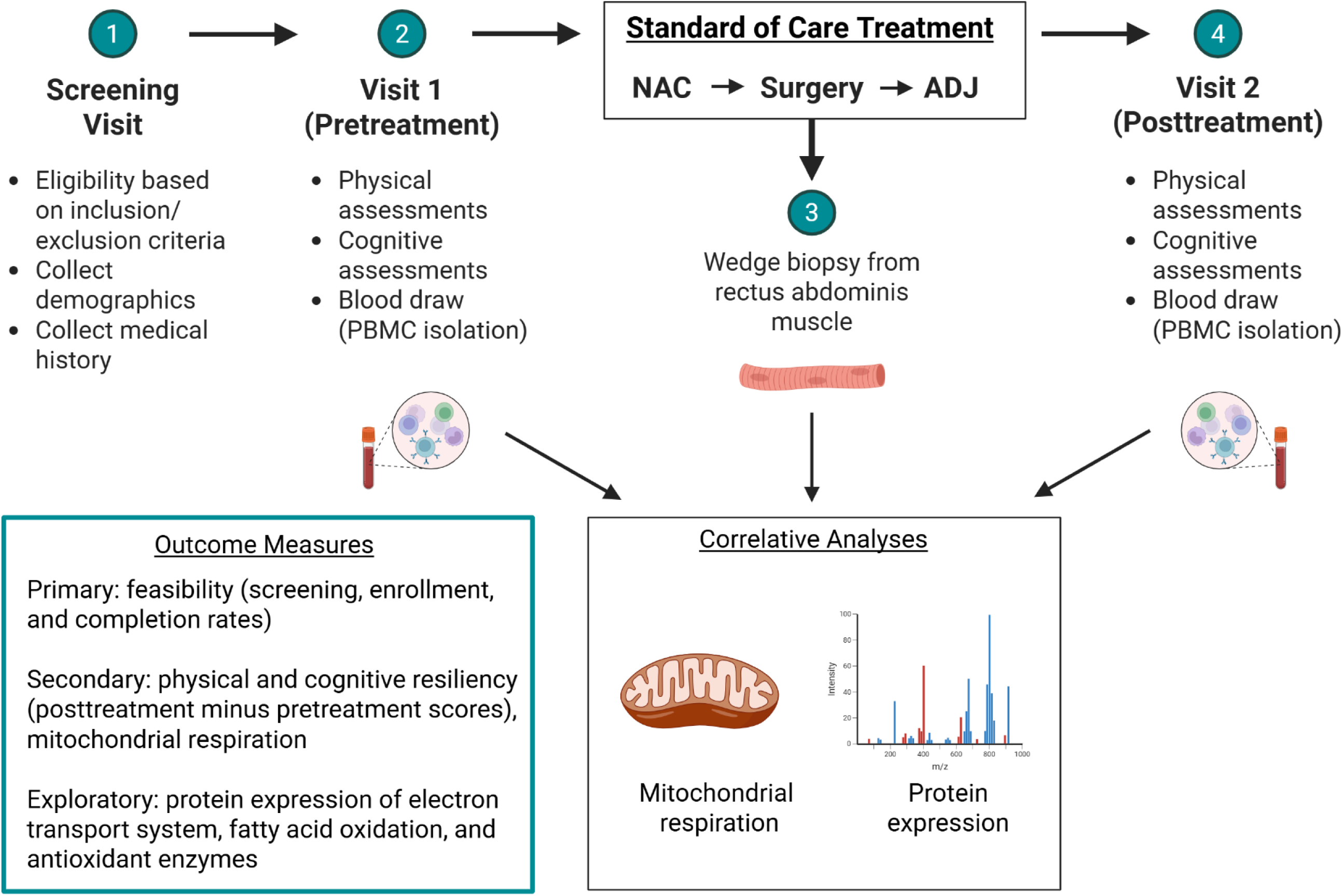
Study overview. *NAC*, neoadjuvant chemotherapy; *ADJ*, adjuvant chemotherapy

Eligible participants are adults aged 50 years and older with newly diagnosed or suspected FIGO Stage II-IV ovarian, primary peritoneal, fallopian tube, or endometrial carcinoma of any histological subtype. Participants planned treatment must include neoadjuvant chemotherapy, interval cytoreductive surgery, and adjuvant chemotherapy. Detailed inclusion and exclusion criteria are shown in Table 1.

**Table 1.** Enrollment criteria.

| Inclusion criteria | Exclusion criteria |
| --- | --- |
| • Age ≥ 50 years<br>• Newly diagnosed or suspected FIGO Stage II-IV ovarian, primary peritoneal, fallopian tube, or endometrial carcinoma of any histological subtype<br>• Planned, standard of care treatment consisting of neoadjuvant chemotherapy, tumor reduction surgery, and adjuvant chemotherapy<br>• Ability to understand and willingness to sign IRB-approved informed consent<br>• Ability to read and understand the English language | • Previously treated for ovarian or endometrial cancer<br>• History of brain metastases, whole brain irradiation, poorly controlled psychiatric disorder defined by hospitalization within the last 3 months for psychiatric diagnoses, traumatic brain injury, cerebrovascular event, or dementia<br>• Currently taking anti-amyloid agents, cholinesterase inhibitors, or glutamate regulator<br>• Documented physical or psychological comorbidity that would limit ability to understand or comply with study procedures for the entire length of the study |

### Procedures

#### Study visits and assessments

Eligible participants who provide written informed consent will attend two study visits (Table 2), the first prior to initiation of neoadjuvant chemotherapy, and the second within 30 days of completion of all front-line standard of care treatment. Participants will undergo objective, standardized physical and cognitive assessments, and patient-reported symptoms will be collected concurrently. Physical assessments include the Short Physical Performance Battery (SPPB) (Guralnik et al., 1994) and measurement of handgrip strength using an isometric hydraulic hand dynamometer (Jamar, Bolingbrook, IL). Cognitive assessments include the Wide Range Achievement Test (WRAT5) - Fifth Edition Word Reading (Wilkinson and Robertson, 2006) to assess reading ability and estimate premorbid intelligence (Strauss et al., 2006), Hopkins Verbal Learning Test-Revised (HVLT-R) (Benedict et al., 1998) to assess verbal learning and memory, Trail-Making Test (TMT) Parts A and B (Spreen and Strauss; Tombaugh, 2004) to assess psychomotor speed and cognitive flexibility, Controlled Oral Word Association Test (COWAT) from the Multilingual Aphasia Examination (MAE) (Benton et al., 1983; Strauss et al., 2006) to assess verbal fluency, and the Neuropsychological Assessment Battery (NAB) Naming Test (Stern and White, 2003; Yochim et al., 2009) to assess confrontation naming. Patient-reported symptoms will be collected using the Edmonton Symptom Assessment System - Revised (ESAS-r). Blood will be drawn at both study visits for correlative studies. Participants are reimbursed after completion of each visit.

**Table 2.**
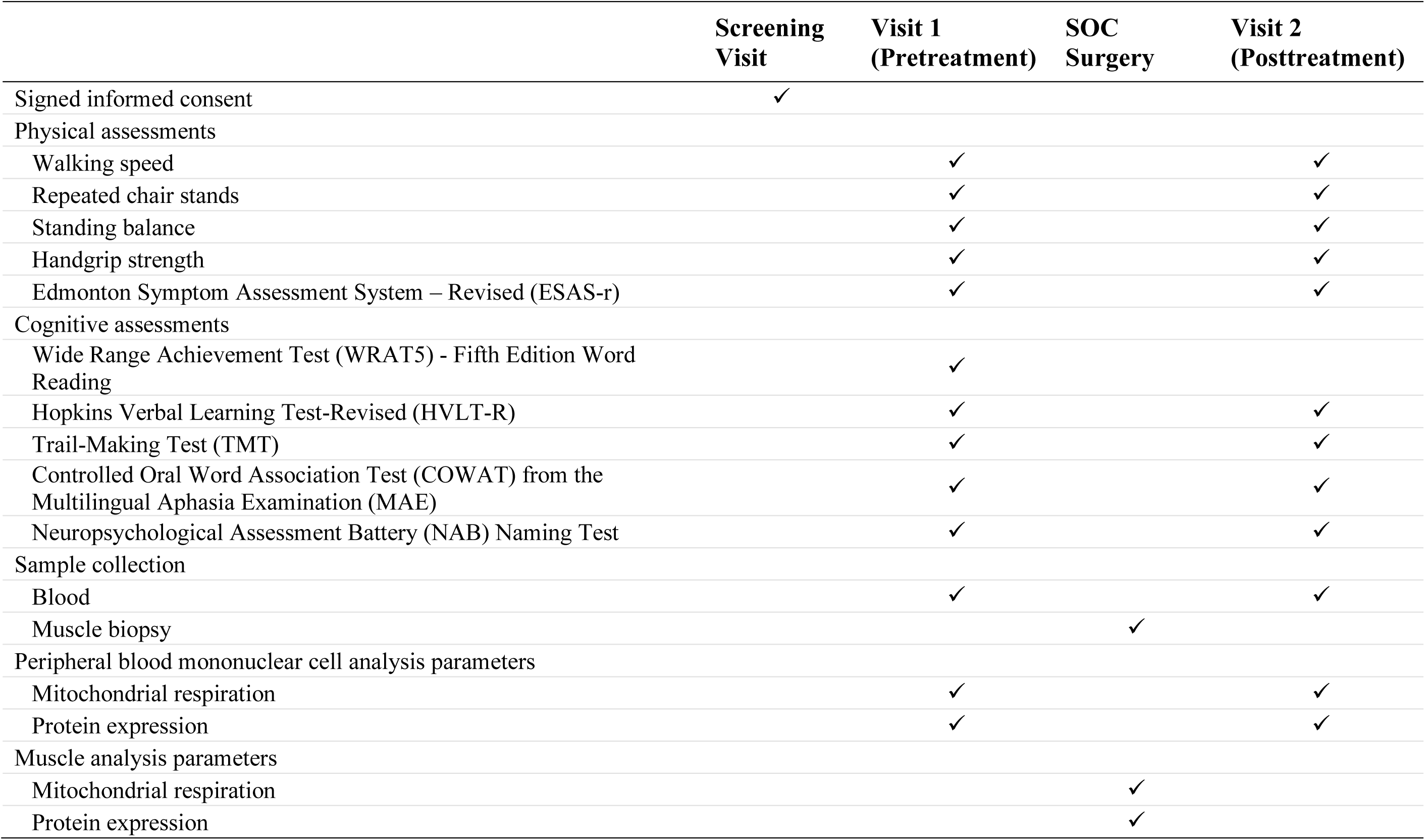
Assessments and visits.

#### Biological sample processing and freezing

During cytoreductive surgery, a skeletal muscle biopsy (∼200-300 mg) from rectus abdominus or psoas muscle will be obtained via wedge biopsy. Muscle tissue will be snap frozen in liquid nitrogen and stored at -80 °C until bioenergetic analysis. Blood samples (∼30 mL per participant) will be collected in EDTA tubes, which will be stored at ambient temperature prior to processing. Peripheral blood mononuclear cells (PBMCs) will be isolated following established methods. Briefly, blood is centrifuged at 500 x g for 15 min, the buffy coat collected and diluted 4x with RPMI 1640 before layering on Histopaque 1077 for density gradient centrifugation at 700 x g for 30 min. The mononuclear cell layer is washed at 500 x g in 25 ml for 15 min, then twice at 400 x g in 4 ml RPMI for 5 min, then counted. The PBMCs will be cryopreserved for assessment of mitochondrial bioenergetics described below.

#### Mitochondrial bioenergetics

Mitochondrial bioenergetics using frozen tissues will be measured following a previously established method (Acin-Perez et al., 2020b). Fig. 2 shows the feasibility of mitochondrial OCR measures using frozen mouse skeletal muscle samples with the following methods. Snap frozen muscle tissues will be thawed in ice-cold PBS, minced, and homogenized using Teflon-glass homogenizer in MAS buffer, containing 220 mM sucrose, 70 mM mannitol, 10 mM K_2_HPO_4_, 5 mM MgCl_2_, 2 mM HEPES, 1 mM EGTA, and 0.2% fatty acid free BSA. Collagenase Type II will be added to this buffer solution at a concentration of 0.25 mg/ml. The samples will be incubated at 37 °C for 30 min. Homogenates will be centrifuged at 1,000 x g for 10 min at 4 °C; then, the supernatant will be collected, and protein concentration will be determined by bicinchoninic acid assay (BCA) (Thermo Fisher). Six ug of protein homogenates in 20 ul of MAS buffer will be loaded into each Seahorse XF96 microplate in triplicates. The loaded plate will be centrifuged at 2,000 x g for 5 min at 4 °C with no brake and an additional 130 ul of MAS supplemented with cytochrome c (10 ug/ml, final concentration) will be added to each well.

**Fig 2.**
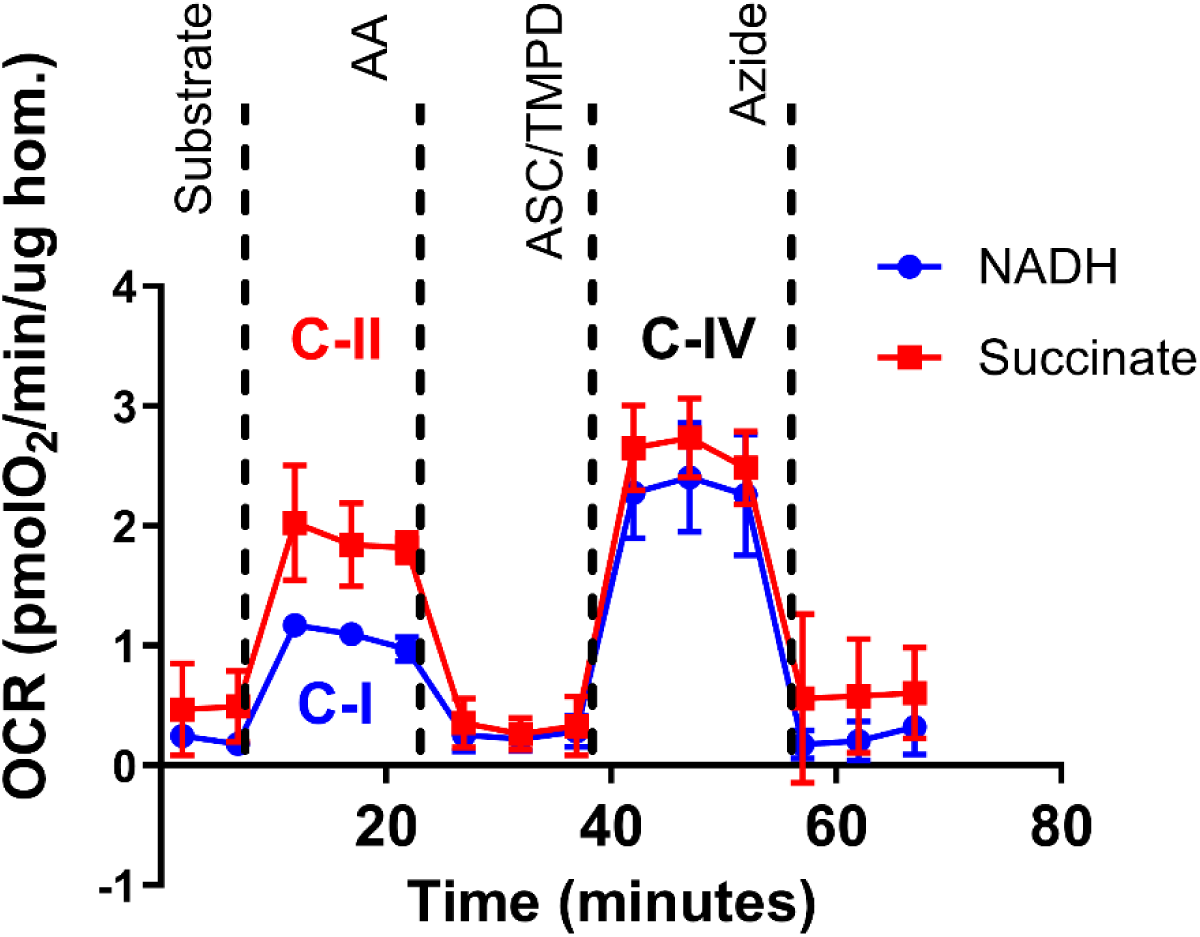
Representative oxygen consumption rate (OCR) traces in frozen mouse gastrocnemius muscle homogenates in response to complex-specific substrates. The blue trace shows OCR following NADH addition, activating complex I (C-I) enzymes. The red trace shows OCR following succinate addition, activating complex II (C-II) enzymes. Ascorbate (ASC) and TMPD were subsequently added to stimulate complex IV (C-IV) activity in both conditions. Antimycin A (AA) was used to inhibit complex III and confirm mitochondrial specificity. *n* = 3. Abbreviations: OCR, oxygen consumption rate; C-I, complex I; C-II, complex II; C-IV, complex IV; AA, antimycin A; ASC, ascorbate.

Substrate injection will be as follows: pyruvate + malate (5 mM each), NADH (1 mM), or 5 mM succinate + rotenone (5 mM + 2 uM) will be injected at port A; rotenone + antimycin A (2 uM + 4 uM) at port B; TMPD + ascorbic acid (0.5 mM + 1 mM) at port C; and azide (50 mM) at port D. These conditions will allow for the determination of the respiratory capacity of mitochondria through complex I, II and IV (Acin-Perez et al., 2020a). PBMCs will be thawed and suspended in XF-DMEM media to seed 300,000 cells per well in 30 ul media. The plate will be centrifuged at 1,300 x g for 1 min with no brake, then rotated by 180 degrees and centrifuged again. After centrifugation, 150 ul of MAS buffer supplemented with cytochrome c (10 uM) and alamethicin (ALA) 1.5 ug/ml will be added in each well. The cartridge will be loaded with the substrates and inhibitors that will activate individual complexes of the mitochondria as described above.

#### Mass spectrometry-based proteomics

Protein expression of electron transport system, fatty acid oxidation, and antioxidant enzymes in muscle tissue and PBMCs will be determined by mass spectrometry (Ahn et al., 2019). PBMC cell pellets will be processed using S-trap micro spin columns (Protifi, Farmingdale, NY) per manufacturer’s protocol. Briefly, all samples will be re-suspended in a uniform volume of 50 μL of lysis buffer containing 5% SDS, 50 mM triethylammonium bicarbonate (TEAB) pH 8.5. After reduction and alkylation, phosphoric acid is then added to a final concentration of 2.5% to obtain a pH ≤ 1, and diluted with 7x volume of binding buffer containing 100 mM TEAB; 90% methanol. The proteins are then trapped on the S-trap columns, washed 3 times with 150 μL of binding buffer, and digested with 20 μL digestion buffer containing 50 mM TEAB and 5 μg of sequencing grade trypsin (Promega, Madison, WI) for 2 h at 47 °C. Eluted peptides will be concentrated by vacuum centrifugation until dry, re-suspended in 20 μL of 0.1% formic acid solution, and quantified using standard BCA protein assay. An aliquot of peptide from each sample will be vialed at a concentration of 0.1 µg/µL and analyzed by LC-MS/MS using data independent acquisition mode (DIA). The MS/MS spectra will be processed using DIA-NN (Demichev et al., 2020) software against the Uniprot human proteome database for peptide identification and quantification. The data will be summarized to the unique protein level and normalized using the median-based central tendency method (Callister et al., 2006).

Muscle tissue will be processed using an integrated workflow that enables measurement of both protein abundance and protein cysteine oxidation (Gluth et al., 2024). Briefly, the biopsies will be minced into small pieces and incubated with lysis buffer containing N-Ethylmaleimide (NEM) to block endogenous free thiols (as specified in (Day et al., 2024)), where the biopsies will be homogenized with a hand-held tissue homogenizer. The lysates will be cleared by centrifugation, protein content in the supernatant determined by BCA, and 200 µg of protein aliquoted for automated cleanup by single-pot, solid-phase–enhanced sample preparation (SP3). The proteins will then be digested using trypsin (Promega) and Lys-C (Wako Chemicals) both at 1:100 ratios for 3 hours and peptides quantified by BCA. Peptide (100 µg) from each sample will be multiplexed using Tandem Mass Tag (TMT18) reagents using a 2.5:1 mass ratio of label:peptide. Following C18 solid phase extraction (SPE) cleanup, the labeled peptides will be quantified by BCA, all reversibly oxidized cysteines will be reduced with DTT, and 5% of total peptide mass will be aliquoted for measuring global protein abundance. The remaining peptide will be used for enrichment of reversibly oxidized cysteines using the resin-assisted capture method (as specified in (Gluth et al., 2024)) and desalted by SPE. Both the global and enriched peptides are then split into 12 fractions each by high-pH reverse phase liquid chromatography (as specified in (Gluth et al., 2024)). Fractionated peptides will be analyzed by LC-MS/MS using a data dependent acquisition mode and the MS/MS spectra searched with MS-GF+ (Kim and Pevzner, 2014) against the Uniprot human proteome database for peptide identification with additional search parameters described elsewhere (Gluth et al., 2024). TMT reporter ion intensities will be extracted by MASIC (Monroe et al., 2008). Peptide identifications will be processed using the “PlexedPiper” R package and workflow (https://github.com/PNNL-Comp-Mass-Spec/PlexedPiper). Within each multiplex, TMT raw reporter ion intensities will be aggregated to unique peptide level and log-2 transformed. Unique peptide level data will be summarized to the protein level to represent the global protein abundance data and will be normalized using the median-based central tendency method (Callister et al., 2006). Unique peptides in the cysteine oxidation data will be summarized to the cysteine site level, where unnormalized unique peptide level global data is used to center the cysteine oxidation data (as specified in (Day et al., 2024)). Both global and cysteine oxidation data will be corrected for multiplex batch effects using the “ComBat” method (Leek et al., 2012). To identify irreversible cysteine oxidation, the PBMCs and muscle global data will also be searched for cysteine dioxidation (sulfinylation) and trioxidation (sulfonylation) modifications and processed in a similar manner as the enriched reversible cysteine oxidation data specified above.

#### Primary outcome measures

The primary objective of this study is to assess the feasibility of performing resilience research in older adults with gynecologic cancers. This will be assessed by calculating screening, enrollment, and study completion rates. The screening rate is the number of approached patients who sign consent divided by the total number of approached patients. The enrollment rate is the number of participants who initiate at least one pretreatment assessment or have pretreatment blood drawn divided by the total number of participants who sign consent. The study completion rate is the number of participants who complete at least one posttreatment assessment (cognitive or physical) divided by the total number of enrollments.

#### Secondary and exploratory outcome measures

For each participant, physical resilience will be measured by the change in score (posttreatment minus pretreatment) for: (1) total ESAS-r; (2) total SPPB; (3) walking speed; (4) repeated chair stands; (5) standing balance; (6) and handgrip strength. Cognitive resilience will be measured by the change in raw and normatively corrected score (posttreatment minus pretreatment) for: (1) HVLT-R total recall, delayed recall, and retention; (2) TMT Part A time (seconds); (3) TMT Part B time (seconds); (4) COWAT total correct; (5) and NAB Naming Test total correct. PBMC and muscle mitochondrial respiration will be measured as the oxygen consumption rate from individual mitochondrial complexes in PBMCs and muscle tissue, respectively. Protein expression of electron transport system, fatty acid oxidation, and antioxidant enzymes will be measured in PBMCs and muscle tissue.

### Statistical methods and considerations

To account for an anticipated 20% withdrawal rate, we will enroll 37 participants to ensure that 30 complete the study. A final sample size of 30 participants will provide 81% power to detect a correlation as low as 0.5 using a two-sided test at a 0.05 significance level. Feasibility will be assessed by calculating participant screening, enrollment, and study completion rates. The study design will be considered feasible if an enrollment rate of at least 80% is achieved. If the observed enrollment rate is 80%, we can be 95% confident that the true enrollment rate lies between 69% and 91%. Similarly, if the final completion rate is 80%, we are 95% confident that the true completion rate will fall between 67%-93%.

For the association analysis for physical functions, multiple linear regressions will be used to examine associations between muscle mitochondrial complex I and II-induced respiration and the changes of physical function (baseline minus posttreatment), a composite score of ESAS-r, SPPB, and grip strength. Models will be adjusted for age, sex, and baseline measures. Similarly, using multiple linear regression models, we will determine the associations between the changes of PBMC mitochondrial complex I and II-induced respiration (baseline minus posttreatment) and changes in cognitive function, a composite score of HVLT-R retention, TMT A and B, COWAT, and NAB Naming Test scores. Again, models will be adjusted for age, sex, and baseline measures. Finally, based on the expression or oxidation levels of key proteins involved in the electron transport system and fatty acid oxidation, we will examine their associations with changes in physical and cognitive resilience, adjusted for age, sex and baseline function measures. Multiple comparisons will be adjusted using false discovery rate (FDR) to ensure an overall FDR < 0.05.

## Discussion

The concept of resilience is novel in the field of aging research and is under-studied in older adults with cancer. In a scoping review of the literature regarding resilience in older adults with cancer, George et al. identified only 29 studies that evaluated physical, cognitive, or psychosocial resilience in adults over age 65 with cancer. The authors identified that the definition of resilience is heterogeneous and highlighted a need for more geroscience-guided studies focused on measuring resilience to identify diagnostic and therapeutic targets to, “positively impact the age-related effects of cancer and cancer treatment in older adults” (George et al., 2023). Additionally, the authors noted that understanding resilience can potentially improve clinical decision-making, such as informing choice of therapy, as well as predicting other outcomes such as survival.

This study will assess the feasibility of conducting resilience research in patients who face numerous other challenges, including the physical, emotional, financial, and logistical challenges of cancer care and management of other comorbidities. Such information will be critical in the design of larger studies to improve our understanding of treatment tolerance in older adults with cancer. To our knowledge, no studies have specifically examined resilience in patients with gynecologic cancers, even though these individuals tend to be older, postmenopausal, and undergo at least two major stressors during the course of treatment, including both chemotherapy and surgery. Resilience to each stressor in part determines the ability of a patient to undergo the next step in treatment (i.e., tolerance of neoadjuvant chemotherapy influences ability to undergo interval cytoreductive surgery; recovery after cytoreductive surgery impacts receipt of adjuvant chemotherapy). Resilience is therefore intrinsically tied to cancer outcomes as completion of both chemotherapy and surgical cytoreduction in the front-line setting has been shown to determine cancer-specific survival (Coleridge et al., 2021).

Our study will also provide new information about the biologic underpinnings of resilience by examining mitochondrial function over time in older adults undergoing cancer treatment with chemotherapy and surgery. Thus far, mitochondrial function has not been well studied as a factor influencing resilience, despite its critical role in human function. Sedrak et al published the largest (to our knowledge) review of measures of aging biology; however, mitochondrial function was not represented. Understanding the mechanisms underlying resilience should help identify opportunities for intervention to improve resilience and decrease vulnerability. Identification of resiliency biomarkers would enable prediction of treatment tolerance as well as monitoring of physiologic responses to interventions (Sedrak et al., 2021).

Our multidisciplinary team uniquely integrates expertise in clinical oncology, neuropsychology, mass spectrometry, and mitochondrial redox biology, enabling a comprehensive approach to studying resilience in cancer patients. This team science approach is critical to developing a comprehensive understanding of the effects of cancer treatment on patients at the phenotypic as well as the molecular level. Our approach represents the future of aging research as it becomes integrated into other areas of medical science and clinical specialties.

## Declarations

Authors declared no competing interests.

## Data Availability

All data produced in the present study are available upon reasonable request to the authors.

